# Musculoskeletal phenotypes in 3q29 deletion syndrome

**DOI:** 10.1101/2023.04.03.23288084

**Authors:** Rebecca M Pollak, Jacob C Tilmon, Melissa M Murphy, Michael J Gambello, Rossana Sanchez Russo, John P Dormans, Jennifer G Mulle

## Abstract

3q29 deletion syndrome (3q29del) is a rare genomic disorder caused by a 1.6 Mb deletion (hg19, chr3:195725000–197350000). 3q29del is associated with neurodevelopmental and psychiatric phenotypes, including an astonishing >40-fold increased risk for schizophrenia, but medical phenotypes are less well-described. We used the online 3q29 registry (3q29deletion.org) to recruit 57 individuals with 3q29del (56.14% male) and requested information about musculoskeletal phenotypes with a custom questionnaire. 85.96% of participants with 3q29del reported at least one musculoskeletal phenotype. Congenital anomalies were most common (70.18%), with pes planus (40.35%), pectus excavatum (22.81%), and pectus carinatum (5.26%) significantly elevated relative to the pediatric general population. 49.12% of participants reported fatigue after 30 minutes or less of activity. Bone fractures (8.77%) were significantly elevated relative to the pediatric general population, suggesting 3q29del impacts bone strength. Participants commonly report receiving medical care for musculoskeletal complaints (71.93%), indicating that these phenotypes impact quality of life for individuals with 3q29del. This is the most comprehensive description of musculoskeletal phenotypes in 3q29del to date, suggests ideas for clinical evaluation, and expands our understanding of the phenotypic spectrum of this syndrome.

## Introduction

The 3q29 deletion is a rare (∼1:30,000) (Kendall et al., 2017; Stefansson et al., 2014) 1.6 Mb deletion on chromosome 3 (hg19, chr3:195725000–197350000) (Ballif et al., 2008; Glassford et al., 2016; Willatt et al., 2005). Individuals with 3q29 deletion syndrome (3q29del) are at substantially increased risk for a variety of neurodevelopmental and neuropsychiatric phenotypes, including mild to moderate intellectual disability (ID), a 19-fold increased risk for autism spectrum disorder (ASD), and a greater than 40-fold increased risk for schizophrenia (SZ) (Ballif et al., 2008; Cox & Butler, 2015; Girirajan et al., 2012; Glassford et al., 2016; Itsara et al., 2009; Kirov et al., 2012; Klaiman et al., 2022; Marshall et al., 2017; Mulle, 2015; Mulle et al., 2010; Pollak et al., 2019; Sanchez Russo et al., 2021; Sanders et al., 2015; Szatkiewicz et al., 2014; Willatt et al., 2005). Medical features of 3q29del are less well-defined, with most reports focusing on the high rate of cardiac manifestations associated with the 3q29 deletion, namely congenital heart defects (Cox & Butler, 2015). However, more work is needed to define the spectrum of other medical phenotypes associated with the 3q29 deletion.

Musculoskeletal complaints, including axial, extremity, and neuromuscular phenotypes, have been associated with an array of copy number variant disorders that have other phenotypic similarities to 3q29del. Significant musculoskeletal phenotypes have been identified in carriers of the 22q11.2 deletion, 16p11.2 reciprocal deletion and duplication, and 7q11.23 deletion (Williams syndrome) (Al-Kateb et al., 2014; Bassett et al., 2005; Chapman et al., 1996; Cherniske et al., 2004; Copes et al., 2016; Damasceno et al., 2014; Digilio et al., 2003; Ficcadenti et al., 2015; Homans et al., 2018; Kaplan et al., 1989; Martin et al., 1984; Morava et al., 2002; Morris et al., 1988; Ren et al., 2020; Ricchetti et al., 2004; Stagi et al., 2010; Taeusch et al., 2004; Wu et al., 2015). These phenotypes include high rates of scoliosis, joint phenotypes including joint laxity and joint contractures, and reduced bone density. Similarly, some musculoskeletal phenotypes have been associated with 3q29del, including chest cavity deformities and finger and toe anomalies (Cox & Butler, 2015; Sanchez Russo et al., 2021). One of these studies, performed by members of our team, diagnosed musculoskeletal anomalies via direct clinical assessment of 3q29 deletion carriers by trained medical geneticists (Sanchez Russo et al., 2021). However, musculoskeletal concerns were not the main focus of either of these studies, and more detail is needed to understand the full phenotypic spectrum associated with the 3q29 deletion.

In the present study, we report the spectrum of musculoskeletal phenotypes systematically ascertained from a large cohort of individuals with 3q29del. Using a custom questionnaire, we were able to assess various domains of musculoskeletal function, including axial, extremity, and neuromuscular phenotypes; imaging procedures; and medical interventions. These data expand our understanding of the greater phenotypic spectrum associated with the 3q29 deletion and provide important insight regarding an under-reported phenotypic domain.

## Methods

### Editorial policies and ethical considerations

This study was approved by Emory University’s Institutional Review Board (IRB00064133) and Rutgers University’s Institutional Review Board (PRO2021001360).

### Sample

57 individuals with 3q29 deletion syndrome (56.14% male) were recruited from the online 3q29 deletion registry (3q29deletion.org), ranging in age from 0.75-46.5 years (mean = 12.02 ± 9.37 years). Inclusion criteria for the present study were membership in the 3q29 deletion registry as of June 2020 and fluency in English. Participants gave informed consent to participate in the study via an online form. A description of the study sample can be found in Table 1.

**Table 1.** Demographics of study participants with 3q29del.

|  | Mean | SD |
| --- | --- | --- |
| **Age** | 12.02 | 9.37 |
|  | N | % |
| **Sex** |  |  |
| Male | 32 | 56.14% |
| Female | 25 | 43.86% |
| **Race** |  |  |
| White | 54 | 94.74% |
| Black | 1 | 1.75% |
| Asian | 2 | 3.51% |
| **Ethnicity** |  |  |
| Not Hispanic/Latino | 40 | 70.18% |
| Hispanic/Latino | 7 | 12.28% |
| Unknown/not reported | 10 | 17.54% |
| **Informant relationship** |  |  |
| Biological parent | 52 | 91.12% |
| Self | 3 | 5.26% |
| Other | 2 | 3.51% |

### Measures

A custom survey instrument was designed by a member of the study team to assess musculoskeletal phenotypes and was reviewed by expert clinicians. The survey consisted of 74 questions across 7 different domains related to musculoskeletal function, including axial and extremity skeleton pathology, joint pathology, neuromuscular function, presence of congenital malformations, other common pediatric musculoskeletal conditions, and any medical treatments received (Supplemental Information). Demographic data was collected separately as a part of the 3q29 deletion registry. Questionnaires were filled out by parents or primary caregivers. The survey was hosted on a local REDCap database, and a link to the survey with instructions was emailed to members of the 3q29 deletion registry between June and July of 2020.

### Analysis

All analyses were performed in R version 4.0.4 (R Core Team, 2008). Data were summarized as counts and proportions of the total participants that reported a given phenotype. The rate of phenotypes in our study sample was compared to the pediatric general population using an exact binomial test via the base R package (R Core Team, 2008). Pediatric general population prevalence rates were established from previously published large studies focusing on pediatric cohorts with an age range similar to that of our study subjects with 3q29del (García-Rodríguez et al., 1999; Konieczny et al., 2013; Naranje et al., 2016; Westphal et al., 2009). Data visualization was performed using the plotly R package (Sievert et al., 2017).

## Results

### High rates of reported musculoskeletal complaints in individuals with 3q29del

Participants with 3q29del overwhelmingly reported musculoskeletal phenotypes, with 85.96% (n=49) of participants reporting at least one musculoskeletal complaint. The most common phenotypic category was congenital anomalies, with 70.18% (n=40) of participants reporting one or more (Figure 1). Excluding congenital phenotypes, 40.35% (n=23) of participants reported at least one joint problem, 15.79% (n=9) of participants reported at least one pain problem, and 59.65% (n=34) of respondents reported at least one neuromuscular problem (Figure 1).

**Figure 1.**
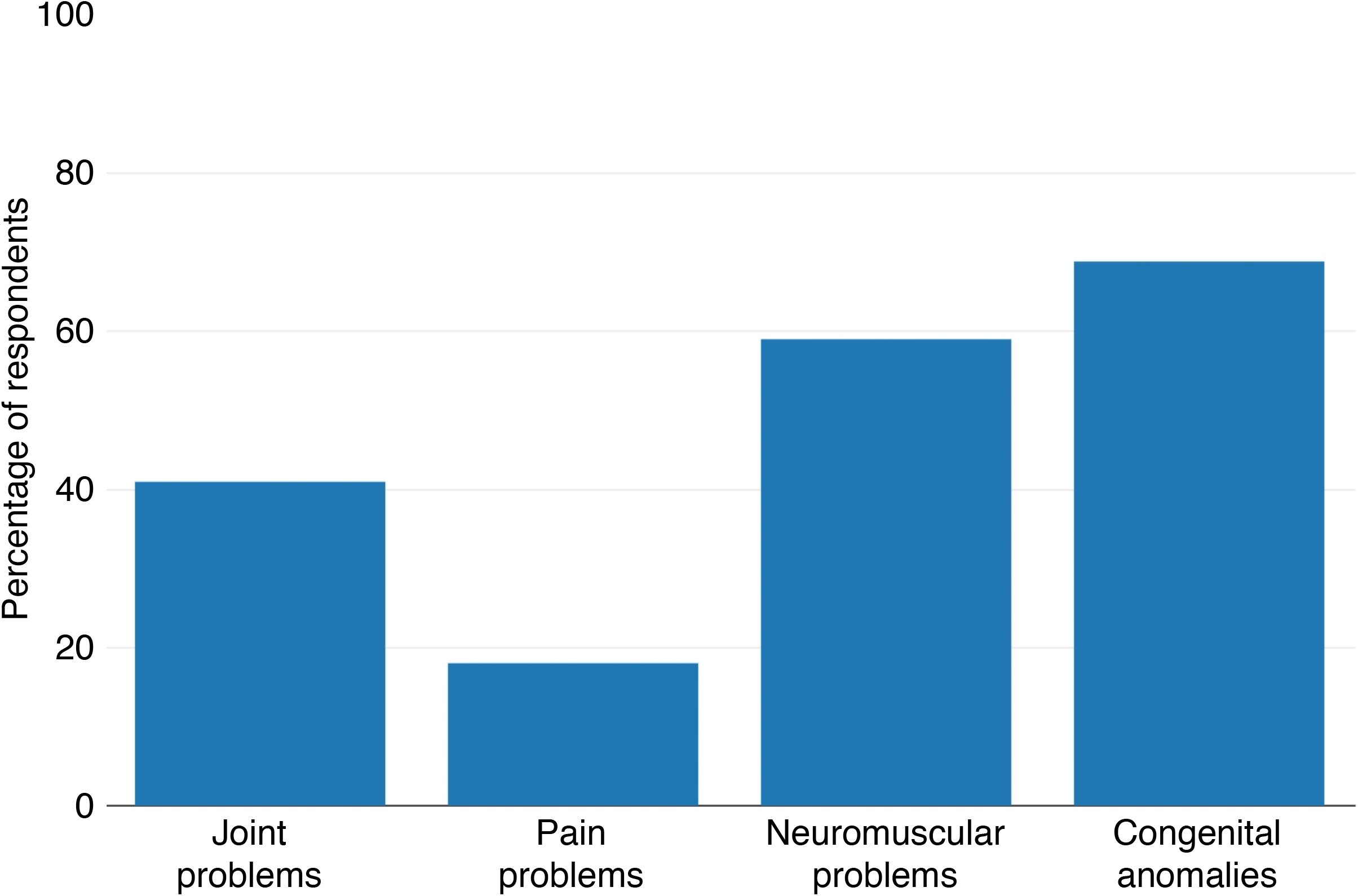
Musculoskeletal phenotypes reported by participants with 3q29del. Rate of musculoskeletal phenotypes across four categories reported by participants with 3q29del (n = 57).

Respondents more commonly reported phenotypes in the extremities (68.42%, n=39) compared to axial (36.84%, n=21) or neuromuscular (59.65%, n=34) manifestations (Table 2). The most commonly reported extremity phenotype was pes planus (40.35%, n=23), followed by joint stiffness (28.07%, n=16) (Table 2). The most commonly reported axial phenotype was pectus excavatum (22.81%, n=13) (Table 2). The most commonly reported neuromuscular phenotype was fatigue in 30 minutes or less of physical activity (49.12%, n=28), followed by gait anomalies (36.84%, n=21) (Table 2).

**Table 2.** Musculoskeletal phenotypes reported by participants with 3q29del.

|  | N | % |
| --- | --- | --- |
| **Axial phenotypes** |  |  |
| Any axial phenotype | 21 | 36.84% |
| Scoliosis | 4 | 7.02% |
| Cervical pain for more than 1 month | 3 | 5.26% |
| Thoracic pain for more than 1 month | 1 | 1.75% |
| Lumbar pain for more than 1 month | 3 | 5.26% |
| Pectus excavatum | 13 | 22.81% |
| Pectus carinatum | 3 | 5.26% |
| Sixth lumbar vertebra | 0 | 0.00% |
| **Extremity phenotypes** |  |  |
| Any extremity phenotype | 39 | 68.42% |
| Joint stiffness | 16 | 28.07% |
| Joint contracture | 2 | 3.51% |
| Joint laxity | 8 | 14.03% |
| Daily joint pain for more than 1 month | 5 | 8.77% |
| Pes planus | 23 | 40.35% |
| Long fingers | 10 | 17.54% |
| Camptodactyly | 2 | 3.51% |
| Nail hypoplasia | 3 | 5.26% |
| Uniquely shaped fingers or toes | 9 | 15.79% |
| Bone fracture | 5 | 8.77% |
| Leg length discrepancy | 0 | 0.00% |
| **Neuromuscular phenotypes** |  |  |
| Any neuromuscular phenotype | 34 | 59.65% |
| Gait anomalies | 21 | 36.84% |
| Fatigue in 30 minutes or less of activity | 28 | 49.12% |

### Variable presence of congenital anomalies within 3q29 deletion carriers

While 68.85% of participants with 3q29del reported at least one congenital anomaly, the specific phenotypes experienced by study participants varied. The most commonly reported congenital anomaly was pes planus (40.35%, n=23), followed by a unique or atypical facial structure (31.58%, n=18) and pectus excavatum (22.81%, n=13) (Figure 2A). To determine whether the reports of congenital anomalies by our study subjects was higher than the pediatric general population, we compared the prevalence of three specific phenotypes. Pes planus (40.35%, n=23, p<2.2E-16), pectus excavatum (22.81%, n=13, p=3.41E-13), and pectus carinatum (5.26%, n=3, p=0.007) were all reported at significantly higher rates by our study subjects compared to the pediatric general population (2.7%, 1.275%, and 0.675%, respectively) (García-Rodríguez et al., 1999; Westphal et al., 2009), suggesting that the 3q29 deletion confers an increased risk for congenital musculoskeletal anomalies.

**Figure 2.**
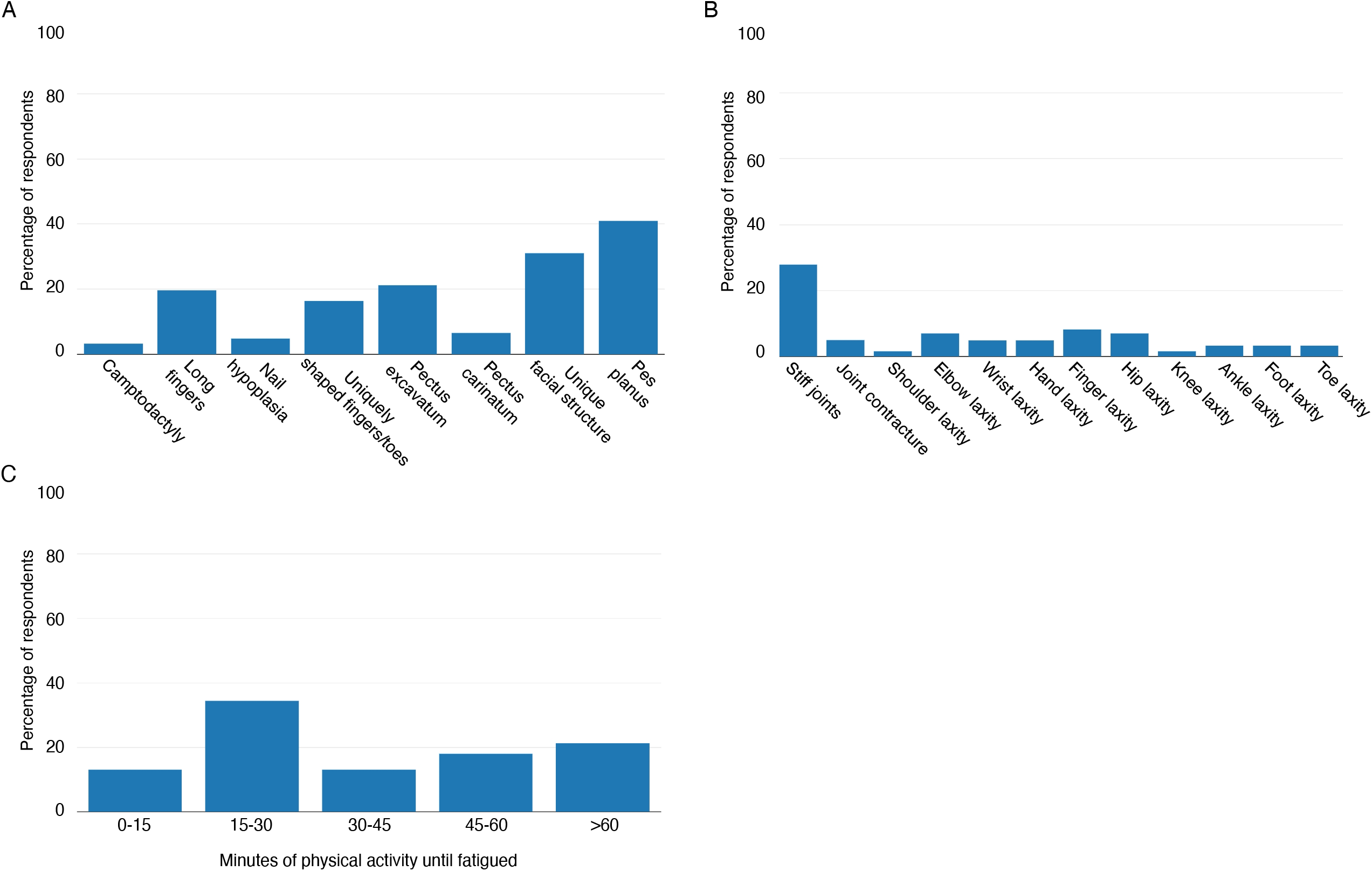
Specific musculoskeletal phenotypes reported by participants with 3q29del. A) Rate of congenital musculoskeletal phenotypes reported by participants with 3q29del (n = 57). B) Rate of joint phenotypes reported by participants with 3q29del (n = 57). C) Amount of physical activity until becoming fatigued reported by participants with 3q29del (n = 57).

### Joint complaints in 3q29 deletion carriers are driven by joint stiffness

Joint problems were a commonly reported phenotype in our study sample. To determine whether there were specific areas of concern versus an equal distribution of complaints across categories, we compared the different classes of joint phenotypes included on our survey instrument. We found that the majority of participants that reported joint problems reported joint stiffness (28.07%, n=16) (Figure 2B). Joint laxity was not commonly reported by our participants, with rates ranging from 0% (shoulder and knee laxity) to 7.02% (n=4, finger laxity) (Figure 2B). These data suggest that the 3q29 deletion does not have a strong phenotypic impact on joint function.

### High rate of fatigue in individuals with 3q29del

To determine the level of musculoskeletal endurance in our study population, we asked how long the individual with 3q29del can participate in physical activity before becoming fatigued. 49.12% (n=28) of our study participants reported becoming fatigued after 30 minutes or less of physical activity (Table 2, Figure 2C). Study participants most commonly reported becoming fatigued after 15 to 30 minutes of physical activity (36.84%, n=21) (Figure 2C). These data suggest that musculoskeletal endurance may be impaired by the 3q29 deletion.

### Orthopedic imaging procedures among 3q29del study subjects

To determine the proportion of our study sample that received any medical imaging procedures, we asked participants to report any X-ray, CT scan, or MRI performed. 36.84% (n=21) of our study sample reported receiving at least one imaging procedure. The most commonly reported imaging procedures were spinal X-ray and skull MRI (12.28%, n=7) (Table 3). Rates of extremity imaging procedures were relatively low; the most commonly reported extremity imaging procedures were hand and hip X-rays (8.77%, n=5) (Table 3). These data suggest that the musculoskeletal phenotypes associated with the 3q29 deletion may not commonly require advanced imaging procedures.

**Table 3.** Imaging procedures reported by participants with 3q29del.

| Body part | Imaging procedure | N | % |
| --- | --- | --- | --- |
| Skull | X-ray | 1 | 1.75% |
|  | CT | 4 | 7.02% |
|  | MRI | 7 | 12.28% |
| Neck | X-ray | 1 | 1.75% |
|  | CT | 0 | 0.00% |
|  | MRI | 3 | 5.26% |
| Spine | X-ray | 7 | 12.28% |
|  | CT | 0 | 0.00% |
|  | MRI | 3 | 5.26% |
| Shoulder | X-ray | 2 | 3.51% |
|  | CT | 0 | 0.00% |
|  | MRI | 0 | 0.00% |
| Arm | X-ray | 2 | 3.51% |
|  | CT | 0 | 0.00% |
|  | MRI | 0 | 0.00% |
| Elbow | X-ray | 0 | 0.00% |
|  | CT | 0 | 0.00% |
|  | MRI | 0 | 0.00% |
| Hand | X-ray | 5 | 8.77% |
|  | CT | 0 | 0.00% |
|  | MRI | 0 | 0.00% |
| Rib | X-ray | 1 | 1.75% |
|  | CT | 0 | 0.00% |
|  | MRI | 0 | 0.00% |
| Pelvis | X-ray | 2 | 3.51% |
|  | CT | 0 | 0.00% |
|  | MRI | 1 | 1.75% |
| Hip | X-ray | 5 | 8.77% |
|  | CT | 0 | 0.00% |
|  | MRI | 0 | 0.00% |
| Leg | X-ray | 1 | 1.75% |
|  | CT | 0 | 0.00% |
|  | MRI | 1 | 1.75% |
| Knee | X-ray | 2 | 3.51% |
|  | CT | 0 | 0.00% |
|  | MRI | 2 | 3.51% |
| Ankle | X-ray | 3 | 5.26% |
|  | CT | 0 | 0.00% |
|  | MRI | 0 | 0.00% |
| Foot | X-ray | 3 | 5.26% |
|  | CT | 0 | 0.00% |
|  | MRI | 0 | 0.00% |

### Elevated rate of bone fractures associated with the 3q29 deletion

Coincident with the relatively low rates of diagnostic imaging received by our study sample, few participants reported medical diagnoses confirmed by imaging procedures. Notably, 8.77% (n=5) of our study sample reported a bone fracture confirmed by diagnostic imaging (Table 4); this is significantly elevated compared to the pediatric general population prevalence of 0.947% (p=0.0002) (Naranje et al., 2016). 7.02% (n=4) of our study sample reported scoliosis confirmed by diagnostic imaging (Table 4); this is not significantly increased compared to the pediatric general population prevalence of 0.47-5.2% (midpoint=2.835%, p=0.07) (Konieczny et al., 2013), however it suggests that the 3q29 deletion may affect risk for scoliosis. These data demonstrate that the 3q29 deletion is associated with bone fracture liability. A larger sample size is needed to definitively determine whether 3q29 deletion participants have scoliosis at a significantly increased rate compared to the pediatric general population.

**Table 4.** Participant-reported musculoskeletal diagnoses confirmed by imaging procedures.

| Diagnosis | N | % |
| --- | --- | --- |
| Bone fracture | 5 | 8.77% |
| Scoliosis | 4 | 7.02% |
| Leg length discrepancy | 0 | 0.00% |
| Mass/tumor | 0 | 0.00% |
| Sixth lumbar vertebra | 0 | 0.00% |
| Other condition | 4 | 7.02% |

### Medical care for musculoskeletal complaints is common for 3q29 deletion carriers

We sought to determine whether the musculoskeletal complaints present in our study sample of individuals with 3q29del are associated with high rates of medical intervention. Therefore, we asked study participants whether they had ever received physical therapy, an orthopedic evaluation, or orthopedic surgery for their musculoskeletal complaints. 71.93% (n=41) of our study participants reported receiving at least one of these medical interventions. A majority of individuals with 3q29del received physical therapy (64.91%, n=37) (Figure 3). 26.79% (n=15) of our study sample received an orthopedic evaluation, and 7.02% (n=4) underwent orthopedic surgery (Figure 3). These data suggest that some musculoskeletal phenotypes associated with the 3q29 deletion are severe enough to require intervention. These data further suggest that musculoskeletal complaints in 3q29 deletion syndrome may be improved or corrected with less invasive medical interventions such as physical therapy, as compared to surgery.

**Figure 3.**
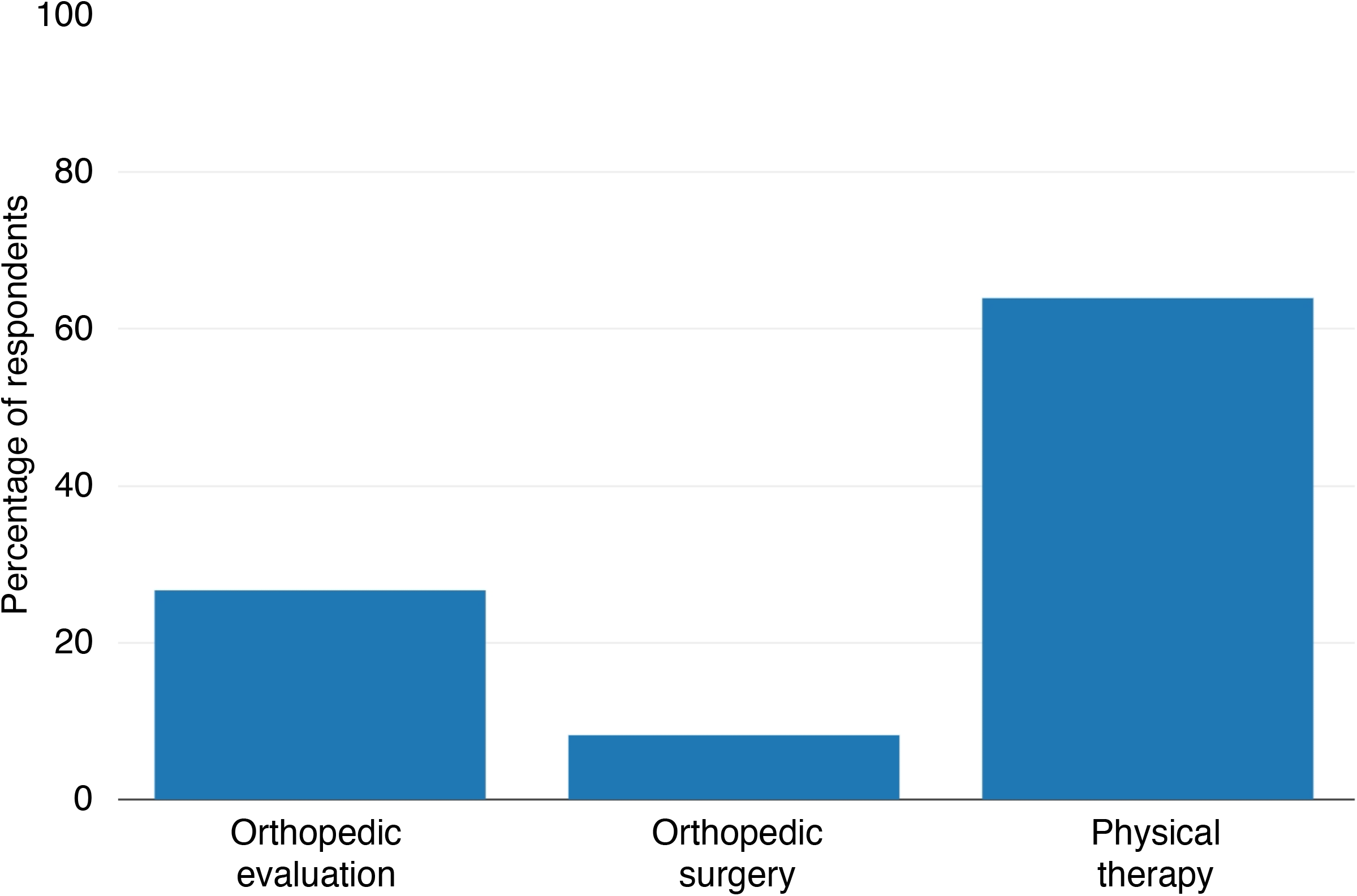
Medical interventions reported by participants with 3q29del. Rate of medical interventions received for musculoskeletal phenotypes reported by participants with 3q29del (n = 57), showing high rates of medical care needed for musculoskeletal concerns.

## Discussion

This is the first study of 3q29del to systematically ascertain musculoskeletal phenotypes as a primary outcome measure. We found high rates of musculoskeletal phenotypes in our study sample of individuals with 3q29del, with 85.96% (n=49) of participants reporting at least one complaint. Congenital anomalies (70.18%, n=40) and neuromuscular problems (59.65%, n=34) were common. Respondents reported more phenotypes localized to the extremities (68.42%, n=39) as compared to the axial skeleton (36.84%, n=21). Individuals with 3q29del also received high rates of medical interventions for their musculoskeletal phenotypes, including physical therapy (64.91%, n=37), orthopedic evaluation (26.79%, n=15), and orthopedic surgery (7.02%, n=4). Together, these data suggest that the 3q29 deletion confers a previously underappreciated risk for musculoskeletal phenotypes.

We found that the rates of several musculoskeletal phenotypes were significantly increased in our study sample relative to the pediatric general population (García-Rodríguez et al., 1999; Naranje et al., 2016; Westphal et al., 2009). Pes planus, pectus excavatum, pectus carinatum, and bone fractures were all elevated in our 3q29del study sample. Scoliosis was not significantly elevated in our study sample relative to the pediatric general population, though there was an increase that is suggestive of an effect (Konieczny et al., 2013). A previous review of case reports and a prior study by our team both identified high rates of chest cavity deformities, and case reports have identified scoliosis in some 3q29 deletion carriers (Cox & Butler, 2015; Sanchez Russo et al., 2021). Together with the present study, these data suggest that the 3q29 deletion confers specific, but underappreciated, risks for musculoskeletal phenotypes.

Musculoskeletal phenotypes have been associated with several copy number variant disorders, including 22q11.2 deletion syndrome, 16p11.2 deletion and duplication syndromes, and Williams syndrome. These syndromes have been associated with scoliosis, extremity anomalies, joint problems, and reduced bone mineral density in isolated studies (Al-Kateb et al., 2014; Bassett et al., 2005; Chapman et al., 1996; Cherniske et al., 2004; Copes et al., 2016; Damasceno et al., 2014; Digilio et al., 2003; Ficcadenti et al., 2015; Homans et al., 2018; Kaplan et al., 1989; Martin et al., 1984; Morava et al., 2002; Morris et al., 1988; Ren et al., 2020; Ricchetti et al., 2004; Stagi et al., 2010; Taeusch et al., 2004; Wu et al., 2015). It has been suggested that the more pressing medical and psychiatric concerns, including congenital heart defects and elevated risk for ASD and SZ, may overshadow orthopedic concerns (Homans et al., 2018). The 3q29 deletion is also associated with a complex constellation of medical, neurodevelopmental, and neuropsychiatric comorbidities that may result in orthopedic phenotypes receiving less attention. However, the high rate of medical care reported by our population specifically for orthopedic concerns highlights the importance of assessing any relevant phenotypes early and introducing appropriate interventions, to lessen the impact of orthopedic problems across the lifespan.

Due to the variable nature of musculoskeletal phenotypes associated with the 3q29 deletion, clinical care should address multiple areas of risk. Based on the present study, in combination with previous reports, individuals with 3q29del should receive regular orthopedic exams, including periodic scoliosis screening (Cox & Butler, 2015; Sanchez Russo et al., 2021). Regularly assessing spinal curvature will allow for early detection of any anomalies, which is critical for maximizing the benefit of early-stage, less-intensive therapeutics, such as bracing. Individuals with 3q29 deletion syndrome should also receive physical therapy assessments, including the six-minute walk test (Andersen et al., 2016; Kuettel et al., 2022), to help guide clinical care. Additionally, the significantly elevated incidence of bone fracture in our study sample suggests that the 3q29 deletion may have an adverse impact on bone mineral density, which may be compounded by documented feeding challenges within the 3q29del population (Wawrzonek et al., 2022). Reduced bone mineral density has also been identified in other copy number variant disorders, including 22q11.2 deletion syndrome and Williams syndrome (Cherniske et al., 2004; Copes et al., 2016; Ficcadenti et al., 2015; Homans et al., 2018; Stagi et al., 2010). To monitor the progression of this phenotype, periodic DEXA scans or another measurement of bone mineral density, paired with basic labs including measurements of calcium and vitamin D levels, will help to guide clinical care. It is important to note the relatively low frequency of medical imaging procedures reported by our study participants; while this may indicate that the musculoskeletal phenotypes in individuals with 3q29del do not commonly require advanced imaging, it may also suggest that musculoskeletal concerns in this population receive less clinical attention as compared to the significant neurodevelopmental and behavioral manifestations of the syndrome. Because our population is relatively young, it is not known whether the 3q29 deletion is a risk factor for late-onset phenotypes related to reduced bone density, such as osteoporosis. These data highlight the importance of longitudinal follow up to assess later onset conditions.

While this study provides critical insight into the musculoskeletal phenotypes associated with 3q29del, it is not without limitations. First, the data was gathered via an optional survey sent out to 3q29 deletion registry participants. It is possible that those individuals who are more severely affected would be more likely to participate. This would potentially bias our results, and lead to an inflation of the rates of different musculoskeletal phenotypes. However, some respondents reported no phenotypes, suggesting that the entire study sample is not comprised of severely affected individuals. Second, the data collected in this study is parent-or self-report, rather than direct clinical observation, and can be subject to recall bias. However, the elevated rates of extremity anomalies, chest wall deformities, and scoliosis mirror a large review of case studies as well as a recent publication by our team that used direct assessment (Cox & Butler, 2015; Sanchez Russo et al., 2021). Our sample is young (average age 12.02 ± 9.37 years) so our study cannot answer questions about later onset phenotypes. Finally, while this is the largest study of musculoskeletal phenotypes in 3q29del to date, the sample size is still limited and lacks racial and ethnic diversity. Future studies will work to increase the enrollment of underrepresented minorities in the online 3q29 deletion registry.

The present study is the first to systematically ascertain musculoskeletal phenotypes in a large sample of individuals with 3q29del. A majority of our study participants reported at least one musculoskeletal phenotype. Congenital phenotypes, including pectus excavatum, pectus carinatum, and pes planus, were frequently reported and were significantly elevated relative to the pediatric general population (García-Rodríguez et al., 1999; Westphal et al., 2009). Participants also reported a high rate of neuromuscular phenotypes, including gait anomalies and fatigue in 30 minutes or less of physical activity. The rate of bone fractures was significantly increased compared to the pediatric general population (Naranje et al., 2016). Medical interventions, particularly physical therapy, were common in our study sample. These data indicate that musculoskeletal phenotypes are a part of the phenotypic spectrum of the 3q29 deletion. Individuals with 3q29del should be monitored for the development of orthopedic concerns, including scoliosis and reduced bone mineral density. Clinical monitoring, combined with early interventions, including physical therapy, where appropriate will improve the function and quality of life for individuals with 3q29del.

## Data Availability

The datasets used and/or analyzed during the current study are available from the corresponding author on reasonable request.

## Acknowledgements

We gratefully acknowledge our study population, the 3q29 deletion community, for their participation and commitment to research.

## Notes

**Competing interests:** The authors have no conflicts of interest to report.

### Competing Interest Statement

The authors have declared no competing interest.

### Funding Statement

This study was funded by NIH R01 MH110701 and NIH T32 GM0008490.

### Author Declarations

Institutional Review Board of Emory University gave ethical approval for this work. Institutional Review Board of Rutgers University gave ethical approval for this work.

## References

Al-Kateb, H., Khanna, G., Filges, I., Hauser, N., Grange, D. K., Shen, J., Smyser, C. D., Kulkarni, S., & Shinawi, M. (2014). Scoliosis and vertebral anomalies: additional abnormal phenotypes associated with chromosome 16p11.2 rearrangement. American Journal of Medical Genetics. Part A, 164a(5), 1118–1126. https://doi.org/10.1002/ajmg.a.36401

Andersen, L. K., Knak, K. L., Witting, N., & Vissing, J. (2016). Two- and 6-minute walk tests assess walking capability equally in neuromuscular diseases. Neurology, 86(5), 442–445. https://doi.org/10.1212/wnl.0000000000002332

Ballif, B. C., Theisen, A., Coppinger, J., Gowans, G. C., Hersh, J. H., Madan-Khetarpal, S., Schmidt, K. R., Tervo, R., Escobar, L. F., Friedrich, C. A., McDonald, M., Campbell, L., Ming, J. E., Zackai, E. H., Bejjani, B. A., & Shaffer, L. G. (2008). Expanding the clinical phenotype of the 3q29 microdeletion syndrome and characterization of the reciprocal microduplication. Molecular Cytogenetics, 1, 8. https://doi.org/10.1186/1755-8166-1-8

Bassett, A. S., Chow, E. W., Husted, J., Weksberg, R., Caluseriu, O., Webb, G. D., & Gatzoulis, M. A. (2005). Clinical features of 78 adults with 22q11 Deletion Syndrome. American Journal of Medical Genetics. Part A, 138(4), 307–313. https://doi.org/10.1002/ajmg.a.30984

Chapman, C. A., du Plessis, A., & Pober, B. R. (1996). Neurologic findings in children and adults with Williams syndrome. J Child Neurol, 11(1), 63–65. https://doi.org/10.1177/088307389601100116

Cherniske, E. M., Carpenter, T. O., Klaiman, C., Young, E., Bregman, J., Insogna, K., Schultz, R. T., & Pober, B. R. (2004). Multisystem study of 20 older adults with Williams syndrome. American Journal of Medical Genetics Part A, 131(3), 255–264.

Copes, L. E., Pober, B. R., & Terilli, C. A. (2016). Description of common musculoskeletal findings in Williams Syndrome and implications for therapies. Clin Anat, 29(5), 578–589. https://doi.org/10.1002/ca.22685

Cox, D. M., & Butler, M. G. (2015). A clinical case report and literature review of the 3q29 microdeletion syndrome. Clinical dysmorphology, 24(3), 89–94. https://doi.org/10.1097/MCD.0000000000000077

Damasceno, M. L., Cristante, A. F., Marcon, R. M., & Barros Filho, T. E. (2014). Prevalence of scoliosis in Williams-Beuren syndrome patients treated at a regional reference center. Clinics (Sao Paulo), 69(7), 452–456. https://doi.org/10.6061/clinics/2014(07)02

Digilio, M. C., Angioni, A., De Santis, M., Lombardo, A., Giannotti, A., Dallapiccola, B., & Marino, B. (2003). Spectrum of clinical variability in familial deletion 22q11.2: from full manifestation to extremely mild clinical anomalies. Clin Genet, 63(4), 308–313. https://doi.org/10.1034/j.1399-0004.2003.00049.x

Ficcadenti, A., Zallocco, F., Neri, R., Giovannini, L., Tirabassi, G., & Balercia, G. (2015). Bone density assessment in a cohort of pediatric patients affected by 22q11DS. J Endocrinol Invest, 38(10), 1093–1098. https://doi.org/10.1007/s40618-015-0295-6

García-Rodríguez, A., Martín-Jiménez, F., Carnero-Varo, M., Gómez-Gracia, E., Gómez-Aracena, J., & Fernández-Crehuet, J. (1999). Flexible flat feet in children: a real problem? Pediatrics, 103(6), e84. https://doi.org/10.1542/peds.103.6.e84

Girirajan, S., Rosenfeld, J. A., Coe, B. P., Parikh, S., Friedman, N., Goldstein, A., Filipink, R. A., McConnell, J. S., Angle, B., & Meschino, W. S. (2012). Phenotypic heterogeneity of genomic disorders and rare copy-number variants. New England Journal of Medicine, 367(14), 1321–1331.

Glassford, M. R., Rosenfeld, J. A., Freedman, A. A., Zwick, M. E., Mulle, J. G., & Unique Rare Chromosome Disorder Support, G. (2016). Novel features of 3q29 deletion syndrome: Results from the 3q29 registry. American Journal of Medical Genetics. Part A, 170A(4), 999–1006. https://doi.org/10.1002/ajmg.a.37537

Homans, J. F., Tromp, I. N., Colo, D., Schlösser, T. P. C., Kruyt, M. C., Deeney, V. F. X., Crowley, T. B., McDonald-McGinn, D. M., & Castelein, R. M. (2018). Orthopaedic manifestations within the 22q11.2 Deletion syndrome: A systematic review. American Journal of Medical Genetics. Part A, 176(10), 2104–2120. https://doi.org/10.1002/ajmg.a.38545

Itsara, A., Cooper, G. M., Baker, C., Girirajan, S., Li, J., Absher, D., Krauss, R. M., Myers, R. M., Ridker, P. M., Chasman, D. I., Mefford, H., Ying, P., Nickerson, D. A., & Eichler, E. E. (2009). Population analysis of large copy number variants and hotspots of human genetic disease. American Journal of Human Genetics, 84(2), 148–161. https://doi.org/10.1016/j.ajhg.2008.12.014

Kaplan, P., Kirschner, M., Watters, G., & Costa, M. T. (1989). Contractures in patients with Williams syndrome. Pediatrics, 84(5), 895–899.

Kendall, K. M., Rees, E., Escott-Price, V., Einon, M., Thomas, R., Hewitt, J., O’Donovan, M. C., Owen, M. J., Walters, J. T. R., & Kirov, G. (2017). Cognitive Performance Among Carriers of Pathogenic Copy Number Variants: Analysis of 152,000 UK Biobank Subjects. Biological Psychiatry, 82(2), 103–110. https://doi.org/10.1016/j.biopsych.2016.08.014

Kirov, G., Pocklington, A. J., Holmans, P., Ivanov, D., Ikeda, M., Ruderfer, D., Moran, J., Chambert, K., Toncheva, D., Georgieva, L., Grozeva, D., Fjodorova, M., Wollerton, R., Rees, E., Nikolov, I., van de Lagemaat, L. N., Bayés, A., Fernandez, E., Olason, P. I., Owen, M. J. (2012). De novo CNV analysis implicates specific abnormalities of postsynaptic signalling complexes in the pathogenesis of schizophrenia. Molecular Psychiatry, 17(2), 142–153. https://doi.org/10.1038/mp.2011.154

Klaiman, C., White, S. P., Saulnier, C., Murphy, M., Burrell, L., Cubells, J., Walker, E., & Mulle, J. G. (2022). A distinct cognitive profile in individuals with 3q29 deletion syndrome. J Intellect Disabil Res. https://doi.org/10.1111/jir.12919

Konieczny, M. R., Senyurt, H., & Krauspe, R. (2013). Epidemiology of adolescent idiopathic scoliosis. J Child Orthop, 7(1), 3–9. https://doi.org/10.1007/s11832-012-0457-4

Kuettel, J., Bay, R. C., & McIsaac, T. L. (2022). Configuration variability of the six-minute walk test among licensed physical therapists working with neurologic conditions: a pilot survey. Physiotherapy Theory and Practice, 1–17. https://doi.org/10.1080/09593985.2022.2140318

Marshall, C. R., Howrigan, D. P., Merico, D., Thiruvahindrapuram, B., Wu, W., Greer, D. S., Antaki, D., Shetty, A., Holmans, P. A., Pinto, D., Gujral, M., Brandler, W. M., Malhotra, D., Wang, Z., Fajarado, K. V. F., Maile, M. S., Ripke, S., Agartz, I., Albus, M., Schizophrenia Working Groups of the Psychiatric Genomics, C. (2017). Contribution of copy number variants to schizophrenia from a genome-wide study of 41,321 subjects. Nature Genetics, 49(1), 27–35. https://doi.org/10.1038/ng.3725

Martin, N., Snodgrass, G., & Cohen, R. (1984). Idiopathic infantile hypercalcaemia--a continuing enigma. Archives of Disease in Childhood, 59(7), 605–613.

Morava, E., Lacassie, Y., King, A., Illes, T., & Marble, M. (2002). Scoliosis in velo-cardio-facial syndrome. J Pediatr Orthop, 22(6), 780–783.

Morris, C. A., Demsey, S. A., Leonard, C. O., Dilts, C., & Blackburn, B. L. (1988). Natural history of Williams syndrome: physical characteristics. The Journal of Pediatrics, 113(2), 318–326.

Mulle, J. G. (2015). The 3q29 deletion confers >40-fold increase in risk for schizophrenia. Molecular Psychiatry, 20(9), 1028–1029. https://doi.org/10.1038/mp.2015.76

Mulle, J. G., Dodd, A. F., McGrath, J. A., Wolyniec, P. S., Mitchell, A. A., Shetty, A. C., Sobreira, N. L., Valle, D., Rudd, M. K., Satten, G., Cutler, D. J., Pulver, A. E., & Warren, S. T. (2010). Microdeletions of 3q29 confer high risk for schizophrenia. American Journal of Human Genetics, 87(2), 229–236. https://doi.org/10.1016/j.ajhg.2010.07.013

Naranje, S. M., Erali, R. A., Warner, W. C., Jr., Sawyer, J. R., & Kelly, D. M. (2016). Epidemiology of Pediatric Fractures Presenting to Emergency Departments in the United States. J Pediatr Orthop, 36(4), e45–48. https://doi.org/10.1097/bpo.0000000000000595

Pollak, R. M., Murphy, M. M., Epstein, M. P., Zwick, M. E., Klaiman, C., Saulnier, C. A., the Emory 3q29 Project, & Mulle, J. G. (2019). Neuropsychiatric phenotypes and a distinct constellation of ASD features in 3q29 deletion syndrome: results from the 3q29 registry. Mol Autism, 10(1), 30. https://doi.org/10.1186/s13229-019-0281-5

R Core Team. (2008). R: A language and environment for statistical computing. R Foundation for Statistical Computing, Vienna, Austria. https://www.r-project.org/

Ren, X., Yang, N., Wu, N., Xu, X., Chen, W., Zhang, L., Li, Y., Du, R. Q., Dong, S., Zhao, S., Chen, S., Jiang, L. P., Wang, L., Zhang, J., Wu, Z., Jin, L., Qiu, G., Lupski, J. R., Shi, J., Liu, P. (2020). Increased TBX6 gene dosages induce congenital cervical vertebral malformations in humans and mice. Journal of Medical Genetics, 57(6), 371–379. https://doi.org/10.1136/jmedgenet-2019-106333

Ricchetti, E. T., States, L., Hosalkar, H. S., Tamai, J., Maisenbacher, M., McDonald-McGinn, D. M., Zackai, E. H., & Drummond, D. S. (2004). Radiographic study of the upper cervical spine in the 22q11.2 deletion syndrome. J Bone Joint Surg Am, 86(8), 1751–1760. https://doi.org/10.2106/00004623-200408000-00020

Sanchez Russo, R., Gambello, M. J., Murphy, M. M., Aberizk, K., Black, E., Burrell, T. L., Carlock, G., Cubells, J. F., Epstein, M. T., Espana, R., Goines, K., Guest, R. M., Klaiman, C., Koh, S., Leslie, E. J., Li, L., Novacek, D. M., Saulnier, C. A., Sefik, E., Mulle, J. G. (2021). Deep phenotyping in 3q29 deletion syndrome: recommendations for clinical care. Genetics in Medicine, 23(5), 872–880. https://doi.org/10.1038/s41436-020-01053-1

Sanders, S. J., He, X., Willsey, A. J., Ercan-Sencicek, A. G., Samocha, K. E., Cicek, A. E., Murtha, M. T., Bal, V. H., Bishop, S. L., Dong, S., Goldberg, A. P., Jinlu, C., Keaney, J. F., 3rd, Klei, L., Mandell, J. D., Moreno-De-Luca, D., Poultney, C. S., Robinson, E. B., Smith, L., State, M. W. (2015). Insights into Autism Spectrum Disorder Genomic Architecture and Biology from 71 Risk Loci. Neuron, 87(6), 1215–1233. https://doi.org/10.1016/j.neuron.2015.09.016

Sievert, C., Parmer, C., Hocking, T., Chamberlain, S., Ram, K., Corvellec, M., & Despouy, P. (2017). plotly: Create Interactive Web Graphics via ‘plotly.js’. R package version 4.6.0. https://CRAN.R-project.org/package=plotly

Stagi, S., Lapi, E., Gambineri, E., Manoni, C., Genuardi, M., Colarusso, G., Conti, C., Chiarelli, F., de Martino, M., & Azzari, C. (2010). Bone density and metabolism in subjects with microdeletion of chromosome 22q11 (del22q11). Eur J Endocrinol, 163(2), 329–337. https://doi.org/10.1530/eje-10-0167

Stefansson, H., Meyer-Lindenberg, A., Steinberg, S., Magnusdottir, B., Morgen, K., Arnarsdottir, S., Bjornsdottir, G., Walters, G. B., Jonsdottir, G. A., Doyle, O. M., Tost, H., Grimm, O., Kristjansdottir, S., Snorrason, H., Davidsdottir, S. R., Gudmundsson, L. J., Jonsson, G. F., Stefansdottir, B., Helgadottir, I., Stefansson, K. (2014). CNVs conferring risk of autism or schizophrenia affect cognition in controls. Nature, 505(7483), 361–366. https://doi.org/10.1038/nature12818

Szatkiewicz, J. P., O’Dushlaine, C., Chen, G., Chambert, K., Moran, J. L., Neale, B. M., Fromer, M., Ruderfer, D., Akterin, S., Bergen, S. E., Kähler, A., Magnusson, P. K. E., Kim, Y., Crowley, J. J., Rees, E., Kirov, G., O’Donovan, M. C., Owen, M. J., Walters, J., Sullivan, P. F. (2014). Copy number variation in schizophrenia in Sweden [Immediate Communication]. Molecular Psychiatry, 19, 762. https://doi.org/10.1038/mp.2014.40

https://www.nature.com/articles/mp201440#supplementary-information

Taeusch, H., Ballard, R., & Gleason, C. (2004). Avery’s Diseases of the Newborn. Saunders.

Wawrzonek, A. J., Sharp, W., Burrell, T. L., Gillespie, S. E., Pollak, R. M., Murphy, M. M., & Mulle, J. G. (2022). Symptoms of Pediatric Feeding Disorders Among Individuals with 3q29 Deletion Syndrome: A Case-Control Study. J Dev Behav Pediatr, 43(3), e170–e178. https://doi.org/10.1097/dbp.0000000000001009

Westphal, F. L., Lima, L. C., Lima Neto, J. C., Chaves, A. R., Santos Júnior, V. L., & Ferreira, B. L. (2009). Prevalence of pectus carinatum and pectus excavatum in students in the city of Manaus, Brazil. J Bras Pneumol, 35(3), 221–226. https://doi.org/10.1590/s1806-37132009000300005

Willatt, L., Cox, J., Barber, J., Cabanas, E. D., Collins, A., Donnai, D., FitzPatrick, D. R., Maher, E., Martin, H., Parnau, J., Pindar, L., Ramsay, J., Shaw-Smith, C., Sistermans, E. A., Tettenborn, M., Trump, D., de Vries, B. B. A., Walker, K., & Raymond, F. L. (2005). 3q29 microdeletion syndrome: clinical and molecular characterization of a new syndrome. American Journal of Human Genetics, 77(1), 154–160. https://doi.org/10.1086/431653

Wu, N., Ming, X., Xiao, J., Wu, Z., Chen, X., Shinawi, M., Shen, Y., Yu, G., Liu, J., Xie, H., Gucev, Z. S., Liu, S., Yang, N., Al-Kateb, H., Chen, J., Zhang, J., Hauser, N., Zhang, T., Tasic, V., Zhang, F. (2015). TBX6 null variants and a common hypomorphic allele in congenital scoliosis. N Engl J Med, 372(4), 341–350. https://doi.org/10.1056/NEJMoa1406829

