## Supplemental Information for "Musculoskeletal phenotypes in 3q29 deletion syndrome"

**Informant Information**

*Question*: “Name of person completing the survey.”

*Format*: open response.

*Question*: “Relation to person with 3q29.”

*Format*: dropdown menu, only one selection possible.

*Answers*:

Biological parent

Step-parent

Grandparent

Self

Other

**Participant Information**

*Question*: “Child’s name.”

*Format*: open response.

*Question*: “Which of the following conditions does your child have?”

*Format*: checkbox, only one answer possible.

*Answers*:

3q29 Microdeletion Syndrome

3q29 Microduplication Syndrome

**Joint Features**

*Question*: “Have any of your child’s joints been stiff with a limited range of motion?”

*Format*: checkbox, only one answer possible.

*Answers*:

Yes

No

*Question*: “If yes, what joints were stiff with limited range of motion?”

*Format*: open response.

*Question*: “Has your doctor ever diagnosed your child with a joint contracture?”

*Format*: checkbox, only one answer possible.

*Answers*:

Yes

No

*Question*: “If yes, what joints were contracted?”

*Format*: open response.

*Question*: “Are any of your child’s joints chronically swollen? If so, which joints?”

*Format*: open response.

*Question*: “Do any of your child’s joints get recurrently swollen with redness or heat? If so, which joints?”

*Format*: open response.

*Question*: “Are any of your child’s joints more flexible than normal (ligament laxity)? (Select all that apply)”

*Format*: matrix, multiple headings with checkbox next to each heading. Multiple answers allowed for each heading.

*Headings*:

Shoulder

Elbow

Wrist

Hand

Fingers

Hip

Knee

Ankle

Foot

Toes

*Answers*:

Right

Left

**Joint Pain**

*Question*: “Has your child ever had daily joint pain lasting longer than one month?”

*Format*: checkbox, only one answer possible.

*Answers*:

Yes

No

*Question*: “If your child had joint pain, then which joints were involved? (Select all that apply)”

*Format*: matrix, multiple headings with checkbox next to each heading. Multiple answers allowed for each heading.

*Headings*:

Shoulder

Elbow

Wrist

Hand

Fingers

Hip

Knee

Ankle

Foot

Toes

*Answers*:

Right

Left

*Question*: “Has your child ever had neck or back pain lasting longer than one month? If so, what part of the back was involved?”

*Format*: checkbox, multiple answers allowed.

*Answers*:

Neck (Cervical)

Between the shoulder blades (Thoracic)

Lower back (Lumbar)

*Question*: “Did a doctor ever give a reason for your child’s joint or back pain?”

*Format*: checkbox, only one answer possible.

*Answers*:

Yes

No

*Question*: “If applicable, what reason did you child’s doctor give for their joint or back pain?”

*Format*: open response.

**Skeletal Imaging**

*Question*: “Has your child ever had an x-ray, CT, or MRI to look at their bones?”

*Format*: checkbox, only one answer possible.

*Answers*:

Yes

No

*Question*: “If yes, which of the following tests has your child undergone? (Select all that apply)”

*Format*: matrix, multiple headings with checkbox next to each heading. Multiple answers allowed for each heading.

*Headings*:

Skull

Neck

Spine

Shoulder

Arm

Elbow

Hand

Ribs

Pelvis

Hip

Leg

Knee

Ankle

Foot

*Answers*:

X-ray

CT

MRI

*Question*: “Were any of the following conditions confirmed by x-ray, CT, or MRI?”

*Format*: checkbox, multiple answers allowed.

*Answers*:

Broken bone (Fracture)

Scoliosis

One leg found to be longer than the other (Leg length discrepancy)

Mass (Tumor)

Sixth lumbar vertebra

Other (Please specify below)

*Question*: “If applicable, what other condition was diagnosed with x-ray, CT, or MRI?”

*Format*: open response.

*Question*: “If applicable, what bones has your child fractured? (Please include only those confirmed by x-ray, CT, or MRI)

*Format*: open response.

*Question*: “If applicable, what area of the back was affected by scoliosis?”

*Format*: checkbox, multiple answers allowed.

*Answers*:

Above the middle of the back (Proximal thoracic)

Middle of the back (Main thoracic)

Below the middle of the back (Thoracolumbar)

*Question*: “If your child had a confirmed leg length discrepancy, how much was the difference?”

*Format*: open response.

*Question*: “If your child had a mass, where was it located?”

*Format*: open response.

*Question*: “If applicable, what diagnosis did the doctor give for the mass?”

*Format*: open response.

**Past Medical History**

*Question*: “Has your child ever been evaluated by a bone and joint (orthopedic) doctor?”

*Format*: checkbox, only one answer possible.

*Answers*:

Yes

No

*Question*: “If applicable, did your child receive any diagnoses from the orthopedic doctor?”

*Format*: open response.

*Question*: “Has your child ever undergone any bone or joint surgeries?”

*Format*: checkbox, only one answer possible.

*Answers*:

Yes

No

*Question*: “If applicable, what bone or joint surgeries has your child undergone?”

*Format*: open response.

*Question*: “Has your child ever undergone physical therapy?”

*Format*: checkbox, only one answer possible.

*Answers*:

Yes

No

*Question*: “If applicable, for what reason did your child undergo physical therapy?”

*Format*: open response.

**Gait**

*Question*: “Does your child have difficulty walking? (Limping, stumbling, unsteadiness, frequent falls)”

*Format*: checkbox, only one answer possible.

*Answers*:

Yes

No

*Question*: “If applicable, what difficulties does your child have with walking?”

*Format*: open response.

*Question*: “If applicable, has your child’s doctor ever provided a reason for your child’s difficulty with walking?”

*Format*: open response.

*Question*: “How many minutes of physical activity can your child undergo without becoming fatigued?”

*Format*: checkbox, only one answer possible.

*Answers*:

0-15 minutes

15-30 minutes

30-45 minutes

45-60 minutes

Greater than 60 minutes

**Additional Features**

*Question*: “Does your child have any of these additional features?”

*Format*: checkbox, multiple answers allowed.

*Answers*:

Fingers that bend at a wide-angle (Comptodactyly)

Long fingers relative to their hand size

Small or absent nails on the hands and feet (nail hypoplasia)

Unique shape of the fingers or toes

A chest that caves in (Pectus excavatum)

A chest that sticks out (Pectus carinatum)

Unique facial structure

Flat foot

*Question*: “If your child has any of the above features, could you go into more depth as to what that feature looks like in your child?”

*Format*: open response.

*Question*: “Please indicate any information regarding your child’s bones, joints, or muscles that are not included in this survey.”

*Format*: open response.

**Photo Uploads (This section is not required. All photos will be de-identified for privacy.)**

*Question*: “If possible, please upload a photo of your child’s hands with palms up on a neutral background. Please do not include any identifying information in this photo.”

*Format*: document upload.

*Question*: “If possible, please upload a photo of your child’s hands with palms down on a neutral background. Please do not include any identifying information in this photo.”

*Format*: document upload.

*Question*: “Additional photos of your child’s hands (optional).”

*Format*: document upload.

*Question*: “Additional photos of your child’s hands (optional).”

*Format*: document upload.

*Question*: “If possible, please upload photos of the tops of your child’s feet with toes visible on a neutral background. Please do not include any identifying information in these photos.”

*Format*: document upload.

*Question*: “If possible, please upload photos of the soles of your child’s feet with toes visible on a neutral background. Please do not include any identifying information in these photos.”

*Format*: document upload.

*Question*: “Additional photos of your child’s feet (optional).”

*Format*: document upload.

*Question*: “Additional photos of your child’s feet (optional).”

*Format*: document upload.
